# Machine Learning Models for Osteoporosis Prediction: A Systematic Review and Meta-Analysis

**DOI:** 10.64898/2026.07.03.26357134

**Authors:** Filipe Ricardo Carvalho, Paulo Jorge Gavaia

**Author notes:** **Corresponding Author:** Filipe Ricardo Carvalho.

## Abstract

**Purpose:** The application of machine learning (ML) to osteoporosis prediction has expanded rapidly, yet no comprehensive meta-analysis has synthesized the discriminative performance of these models across all ML categories, data types, and validation strategies. This systematic review and meta-analysis aimed to evaluate the diagnostic and predictive accuracy of ML and deep learning models for osteoporosis prediction in adult populations.

**Methods:** Systematic searches of PubMed, Embase, Web of Science, and IEEE Xplore were conducted for studies published between January 2020 and February 2026. Studies developing, validating, or applying ML models for predicting osteoporosis, low bone mineral density, or osteoporotic fractures in adults were included. Methodological quality was assessed using the Prediction Model Risk of Bias Assessment Tool (PROBAST). Area under the receiver operating characteristic curve (AUC) values were pooled using random-effects meta-analysis with logit transformation. Subgroup analyses were performed by data type, ML category, external validation status, and population type. The review followed PRISMA 2020 guidelines.

**Results:** Thirty-three studies were included in the qualitative synthesis and 27 in the meta-analysis. The pooled AUC was 0.879 (95% CI: 0.853–0.901), with substantial heterogeneity (I² = 99.5%). Imaging-based models outperformed clinical data models (AUC = 0.905 vs. 0.872). Deep learning achieved the highest pooled AUC (0.909), followed by ensemble methods (0.874) and traditional ML (0.840). Externally validated models showed lower performance than internally validated ones (AUC = 0.868 vs. 0.897). PROBAST assessment rated 32 of 33 studies (97.0%) as low risk of bias, though this proportion should be interpreted cautiously given that PROBAST was designed for traditional prediction models and may not fully capture ML-specific sources of bias. Egger’s test indicated significant publication bias (p < 0.001). Explainable AI methods were employed in 60.6% of studies, identifying age, body weight, and alkaline phosphatase as the most frequent top predictive features.

**Conclusions:** Machine learning models demonstrate overall good discriminative performance for osteoporosis prediction, albeit with substantial heterogeneity across studies (I² = 99.5%), and show potential as complementary screening tools, particularly in settings with limited DXA access. Deep learning models applied to imaging data and ensemble methods using clinical variables achieved the strongest subgroup estimates. However, extreme heterogeneity, evidence of publication bias, and limited prospective validation warrant cautious interpretation of the pooled estimate. Future research should prioritise multi-centre external validation, standardised reporting following TRIPOD+AI guidelines, and prospective clinical trials to establish real-world clinical impact.

**Mini-Abstract:** This meta-analysis of 33 studies demonstrates that ML models achieve a pooled AUC of 0.879 for osteoporosis prediction, with imaging-based deep learning models reaching 0.905, supporting their potential as complementary screening tools to DXA and FRAX®.

## INTRODUCTION

Osteoporosis is a systemic skeletal disorder characterized by reduced bone mineral density (BMD) and deterioration of bone microarchitecture, leading to increased bone fragility and a heightened susceptibility to fragility fractures—fractures resulting from low-energy trauma equivalent to a fall from standing height or less [1]. It represents a major public health challenge worldwide, affecting an estimated 200 million individuals globally [2], with prevalence rates expected to rise substantially as populations age. Osteoporotic fractures, particularly of the hip, vertebrae, and distal radius, are associated with significant morbidity, mortality, and healthcare costs [2]. Hip fractures alone carry a 12-month mortality rate of approximately 20–30%, and survivors frequently experience lasting functional impairment and loss of independence [3]. The economic burden of osteoporotic fractures is considerable, with annual direct costs exceeding €37 billion in the European Union and $19 billion in the United States [4].

The current gold standard for osteoporosis diagnosis is dual-energy X-ray absorptiometry (DXA), which measures bone mineral density (BMD) at three skeletal sites: the lumbar spine (L1–L4), the proximal femur (femoral neck, trochanter, and total hip), and the one-third (33%) radius. The World Health Organization defines osteoporosis as a T-score of −2.5 or below at these sites, with osteopenia (low bone mass) defined as a T-score between −1.0 and −2.5.[1] However, the principal limitation of DXA as a population-level screening tool is its modest sensitivity for identifying individuals who will sustain an osteoporotic fracture, which has been estimated at below 50%. This limited sensitivity arises because areal BMD measured by DXA captures only one of several determinants of bone strength; in the landmark National Osteoporosis Risk Assessment (NORA) study of over 200,000 post-menopausal women, only a minority of women who subsequently sustained a fragility fracture had a T-score in the osteoporotic range at the time of measurement, while the majority of incident fractures occurred in women with osteopenia or normal BMD [5,6]. In addition, DXA is subject to several practical constraints: the equipment is expensive, requires trained technicians, and is predominantly available in secondary and tertiary care settings [1]. Consequently, DXA access remains uneven, particularly in low- and middle-income countries, rural areas, and primary care settings, contributing to the substantial underdiagnosis of osteoporosis worldwide. Studies have estimated that up to 80% of individuals who have sustained an osteoporotic fracture have not been diagnosed with osteoporosis or received appropriate treatment [7].

Complementary imaging-based assessments have been developed to capture aspects of bone strength not reflected in areal BMD alone. Quantitative computed tomography (QCT) provides volumetric BMD and compartment-specific measurements of cortical and trabecular bone, while trabecular bone score (TBS) derived from lumbar DXA images provides an indirect index of trabecular microarchitecture; both have been incorporated into refined fracture-risk algorithms, including the FRAXplus® extension [8]. Hip structural analysis (HSA), bone turnover markers, and high-resolution peripheral QCT (HR-pQCT) further extend the diagnostic toolkit but remain confined to specialised centres.

To address the limitations of DXA-based screening, several clinical risk assessment tools have been developed to estimate fracture probability using readily available clinical variables. The Fracture Risk Assessment Tool (FRAX®) was developed at the University of Sheffield, then a WHO Collaborating Centre for Metabolic Bone Diseases, and is the most widely used instrument; it calculates the 10-year probability of major osteoporotic and hip fractures from ten clinical risk factors—age, sex, body mass index, prior fragility fracture, parental hip fracture, current smoking, glucocorticoid use, rheumatoid arthritis, secondary osteoporosis, and alcohol intake (≥3 units/day)—with femoral neck BMD as an optional input. FRAX® is currently available in over 80 country-specific calibrations to reflect inter-population variation in fracture incidence and competing mortality, and national guidelines define intervention thresholds in different ways: the National Osteoporosis Guideline Group (NOGG) in the United Kingdom adopts age-dependent assessment and intervention thresholds derived from the 10-year MOF probability of a woman with a prior fragility fracture, whereas the National Osteoporosis Foundation (NOF) in the United States retains fixed thresholds of 20% for major osteoporotic fracture (MOF) and 3% for hip fracture [9,10]. Other tools include the Osteoporosis Self-Assessment Tool (OST), QFracture, and the Garvan Fracture Risk Calculator [11]. While these instruments have demonstrated moderate discriminative ability, they have recognised limitations. The original FRAX® algorithm does not capture the dose–response of certain risk factors, fall history, recency of fracture, type 2 diabetes, or other comorbidities, and its calibration varies across populations and ethnic groups. A recent extension, FRAXplus®, has been developed to incorporate dose–response effects for glucocorticoid use, fall history, recency of prior fracture, trabecular bone score, hip axis length, and other modifiers that were not handled by the original model [12,13]. These limitations have motivated the search for more accurate and adaptable prediction approaches.

Machine learning (ML) and artificial intelligence (AI) have emerged as promising approaches for medical prediction and classification tasks, offering the potential to identify complex, non-linear patterns in high-dimensional data that may be missed by traditional statistical methods. In the field of osteoporosis, ML models have been applied to a range of tasks including the prediction of osteoporosis from clinical and demographic variables, the identification of low BMD from routine radiographs, and the estimation of fracture risk from electronic health records. These models can leverage diverse data sources, including clinical parameters, laboratory values, medical imaging, and increasingly genomic data, and have the flexibility to incorporate hundreds of predictor variables simultaneously. The potential of ML-based approaches is particularly relevant to osteoporosis screening, where the ability to identify high-risk individuals from routinely collected clinical data could enable early intervention and targeted referral for confirmatory DXA scanning.

A wide spectrum of ML algorithms has been applied to osteoporosis prediction [1,7], ranging from traditional approaches such as logistic regression, support vector machines, and random forests to more complex methods including gradient boosting machines (XGBoost, LightGBM, CatBoost), artificial neural networks, and deep learning architectures, such as convolutional neural networks (CNNs). Ensemble methods, which combine predictions from multiple base learners, have been particularly successful in tabular clinical data settings, while deep learning models have shown strong performance in imaging-based tasks such as opportunistic osteoporosis screening from chest radiographs or computed tomography scans. The increasing adoption of explainable AI (XAI) techniques, including SHAP (Shapley Additive Explanations) and LIME (Local Interpretable Model-agnostic Explanations), has begun to address the “black box” concern traditionally associated with complex ML models, facilitating clinical interpretability and trust [14].

The growing body of literature on ML for osteoporosis prediction has prompted several narrative and systematic reviews. Earlier reviews, including those by Smets et al. [15] and Amani et al. [16], provided valuable overviews of the field but were limited by smaller study pools, restricted to specific ML categories or data modalities, or did not include meta-analytic pooling of performance metrics. A subsequent and more comprehensive meta-analysis by Gao et al. [17] pooled the diagnostic performance of artificial-intelligence models for osteoporosis across a broader range of imaging modalities, but predates the rapid expansion of deep-learning and ensemble methods reported in 2022–2025 and did not include studies applying ML to routinely available clinical or laboratory data; the present review therefore extends this body of evidence. Furthermore, the field has evolved rapidly since these reviews were conducted, with a substantial number of new studies published in 2024 and 2025 that incorporate larger datasets, improved validation practices, and novel methodological approaches. To date, no comprehensive systematic review has synthesized the full breadth of recent evidence across all ML categories, data types, and validation strategies while also providing meta-analytic estimates of pooled discriminative performance.

The present systematic review and meta-analysis aim to address this gap by providing a comprehensive and up-to-date synthesis [1,7] of machine-learning and deep-learning models for osteoporosis prediction, classification, and screening. Throughout this review the term “osteoporosis prediction” is used as an umbrella for any model whose target outcome is (i) osteoporosis as defined by the WHO criterion (T-score ≤ −2.5), (ii) low BMD (T-score ≤ −1.0, encompassing osteopenia and osteoporosis), (iii) incident or prevalent osteoporotic fracture, or (iv) a continuous BMD or T-score modelled as a regression task; the specific outcome adopted by each included study is reported in Table 1 and the corresponding results are stratified accordingly in the meta-analysis. Specifically, the objectives were: (1) to systematically identify and characterize all studies published between January 2020 and February 2026 that developed, validated, or applied ML models for osteoporosis prediction in adult populations; (2) to evaluate the discriminative performance of these models through meta-analytic pooling of area under the receiver operating characteristic curve (AUC) [18] values; (3) to conduct subgroup analyses by data type (clinical vs. imaging), ML category (deep learning, ensemble, traditional ML), external validation status, and population type to identify sources of performance variation; (4) to assess methodological quality using the Prediction Model Risk of Bias Assessment Tool (PROBAST); and (5) to identify research gaps and provide recommendations for future studies.

## METHODS

### Protocol and Registration

This systematic review and meta-analysis was conducted in accordance with the Preferred Reporting Items for Systematic Reviews and Meta-Analyses (PRISMA) 2020 guidelines [19]. The review protocol was developed a priori and followed the recommendations of the Cochrane Handbook for Systematic Reviews of Diagnostic Test Accuracy. The study focused on machine learning and deep learning models for the prediction, classification, or screening of osteoporosis, low bone mineral density, or osteoporotic fractures in adult populations.

### Search Strategy

Systematic literature searches were conducted in four electronic databases: PubMed, Embase, Web of Science, and IEEE Xplore, for articles published between January 2020 and February 2026. The lower temporal limit of January 2020 was chosen to capture the rapid expansion of machine-learning research on osteoporosis that followed the wider availability of large clinical imaging datasets and the publication of the PROBAST tool (2019) and the early prediction-model reporting guidance that informed subsequent studies; this window also captures the transition from predominantly classical machine-learning approaches to deep-learning architectures applied to musculoskeletal imaging. Twelve search queries were used combining terms related to machine learning, artificial intelligence, deep learning, and specific algorithms (e.g., XGBoost, random forest, neural network) with terms related to osteoporosis, bone mineral density, and fracture risk prediction. The searches retrieved 298 records from PubMed, 201 from Embase, 87 from Web of Science, and 56 from IEEE Xplore, yielding 642 records in total. After removal of 200 duplicates, 442 unique records were identified. One additional record was identified through manual searching of reference lists, yielding a total of 443 records for screening. The full search strategy for all four databases is provided in the Supplementary Material (Supplementary Search Strategy).

### Eligibility Criteria

#### Inclusion Criteria

Studies were eligible for inclusion if they met all of the following criteria: (1) the study population comprised adults aged 18 years or older at risk of or being screened for osteoporosis; (2) the study developed, validated, or applied a machine learning or deep learning model for predicting or classifying osteoporosis, low bone mineral density (BMD), or osteoporotic fracture risk; (3) the primary outcome corresponded to one of the following: (i) osteoporosis defined by the World Health Organization criterion (femoral neck or lumbar spine T-score ≤ −2.5); (ii) low BMD defined as a T-score ≤ −1.0 (encompassing both osteopenia—T-score between −2.5 and −1.0—and osteoporosis); (iii) incident or prevalent osteoporotic fracture; or (iv) a continuous BMD or T-score outcome modelled as a regression task (hereafter referred to as “BMD regression”); (4) the study design was original research, including cohort, cross-sectional, or case-control studies; (5) the article was published in English; (6) the article was published between January 2020 and February 2026; and (7) at least one performance metric was reported, such as the area under the receiver operating characteristic curve (AUC), sensitivity, specificity, accuracy, or F1 score.

### Exclusion Criteria

Studies were excluded if they were review articles, meta-analyses, editorials, commentaries, or letters to the editor; conference abstracts without full methodology; animal or in vitro studies; studies predicting treatment response rather than osteoporosis diagnosis or risk; studies not focused on osteoporosis (e.g., osteoarthritis classification); studies using only traditional statistical methods without machine learning; or duplicate publications.

### Study Selection Process

The study selection process followed four sequential stages. In Stage 1 (Screening), all 443 records were screened at the title and abstract level against the eligibility criteria. Records clearly not meeting the inclusion criteria were excluded. In Stage 2 (Priority Scoring), the remaining 247 records were assessed for priority based on relevance to machine learning osteoporosis prediction, study design quality, sample size, and publication recency. The 75 highest-priority records were selected for full-text retrieval. This prioritisation step was adopted to ensure feasibility given the large volume of potentially eligible records and to focus resources on studies most directly relevant to machine learning-based osteoporosis prediction; however, it may have introduced selection bias by excluding records that were not identifiable as high-priority at the title and abstract level.

In Stage 3 (Full-Text Eligibility), full texts were sought for all 75 selected records through institutional subscriptions, open-access channels, and inter-library document delivery; for records that remained unavailable, attempts were made to contact the corresponding authors directly. Despite these efforts, nine full texts could not be obtained within the project timeframe and were therefore excluded; the implications of this omission for the completeness of the review are discussed in the Limitations section. The remaining 66 full-text articles were assessed against the eligibility criteria, and 26 were excluded for specific reasons (detailed in Results). Four duplicate file versions of the same study were identified and removed. In Stage 4 (Quality Assessment), the 36 unique studies were subjected to PROBAST risk of bias assessment, during which three additional studies were excluded for fundamental scope or methodological issues.

### Data Extraction

Data were extracted from the included studies using a standardized extraction form comprising 67 fields organized into eight domains: study identification (study ID, first author, year, title, journal, DOI, country, study design, data source), population characteristics (total sample size, training set size, validation set size, external validation set size, age, sex distribution, population type, clinical setting, osteoporosis prevalence in the study sample as defined by the original study [i.e., proportion of participants meeting the study’s own definition of the positive outcome—osteoporosis by WHO T-score, low BMD, prior osteoporotic fracture, or other]), machine learning methodology (all algorithms tested, best-performing algorithm, ML category, feature selection method, number of input features, number of final features, train–test split ratio, cross-validation strategy, hyperparameter tuning, class imbalance handling, validation type), predictor variables (use of DXA, imaging, clinical, or laboratory data as inputs, top predictive features, interpretability method), outcome definition (outcome definition, outcome type, T-score threshold, reference standard, DXA measurement site), performance metrics (AUC, 95% confidence interval, sensitivity, specificity, accuracy, precision, negative predictive value, F1 score, correlation coefficient, mean squared error or root mean squared error, calibration), comparison with existing tools (comparison with FRAX®, FRAX® AUC, comparison with other tools, ML superiority), and quality assessment (PROBAST overall rating and individual domain ratings, external validation status, code availability).

Data were extracted exactly as reported in each study. When a value was not reported, it was recorded as “NR” (not reported). For studies reporting AUC, the performance of the best model on the test or validation set was recorded, rather than training set performance. When multiple outcomes were assessed (e.g., lumbar spine and femoral neck), both values were noted. All machine learning algorithms tested in each study were recorded, not only the best-performing model.

### Quality Assessment

Risk of bias was assessed using the Prediction model Risk Of Bias ASsessment Tool (PROBAST) [20], which evaluates prediction models across four domains: participants, predictors, outcome, and analysis. Each domain was rated as low risk, high risk, or unclear risk of bias. The overall risk of bias for each study was determined by the highest risk rating across any domain. An initial automated assessment was conducted for all 36 studies, followed by manual validation of all studies classified as high or unclear risk of bias (n = 7). During manual validation, studies were reclassified based on detailed review of the full-text articles, and studies with fundamental scope or methodological issues were excluded from the review.

### Data Synthesis and Statistical Analysis

Study characteristics, machine learning methods, and performance metrics were summarized descriptively using frequencies, percentages, medians, and ranges. Studies were categorized by data type (clinical/mixed vs. imaging), machine learning category (deep learning, ensemble methods, traditional ML), validation type (internal only vs. external), and population type (general, postmenopausal, elderly, disease-specific) to facilitate comparison. A random-effects meta-analysis was conducted to estimate the pooled AUC across classification studies. AUC values were logit-transformed prior to pooling to ensure appropriate statistical properties. Standard errors were derived from reported 95% confidence intervals where available; for studies without confidence intervals, standard errors were approximated using the Hanley–McNeil method based on the AUC value and sample size. The DerSimonian–Laird estimator was used for the random-effects model. Back-transformation from the logit scale was applied to obtain the pooled AUC and its 95% confidence interval.

Heterogeneity was assessed using Cochran’s Q statistic and the I² index, where I² values of 25%, 50%, and 75% were interpreted as low, moderate, and high heterogeneity, respectively. Subgroup analyses were performed stratified by data type, machine learning category, external validation status, and population type to explore sources of heterogeneity. Publication bias was assessed using Egger’s regression test and visual inspection of funnel plots. All statistical analyses were performed using Python 3.11 with the SciPy, NumPy, and Matplotlib libraries.

Methodological choice of the summary measure was guided by the Cochrane Handbook for Systematic Reviews of Diagnostic Test Accuracy and the Critical Appraisal and Data Extraction for Systematic Reviews of Prediction Modelling Studies (CHARMS) checklist [21]. We considered the bivariate random-effects model of Reitsma et al. [22] and the hierarchical summary receiver operating characteristic (HSROC) model of Rutter and Gatsonis [23] as alternatives to AUC pooling; however, these approaches require paired sensitivity and specificity values at a defined operating threshold, which were inconsistently reported across the included studies and were defined at study-specific (often unstated) decision cut-offs. AUC pooling was therefore retained as the primary quantitative summary, with the explicit caveat that it is threshold-independent and does not capture trade-offs at clinically relevant operating points. Reporting was further informed by the recently published TRIPOD+AI extension to the TRIPOD statement [24], which provides specific recommendations for prediction-model studies incorporating artificial-intelligence components.

Although the Youden Index (J = sensitivity + specificity − 1) is a useful single-threshold summary of diagnostic test performance and has been applied in several primary studies of AI-based osteoporosis screening, it was not adopted as a pooled summary measure in the present meta-analysis. The principal reason is that the operating thresholds at which sensitivity and specificity were reported differed across the included studies (and were frequently not stated explicitly), so a pooled Youden estimate would have combined non-comparable thresholds. Where reported, study-level sensitivity and specificity are summarised descriptively in the Results.

### Use of Artificial Intelligence

During the preparation of this manuscript, the authors used artificial intelligence writing assistance tools (Claude, Anthropic) to support literature organisation, text drafting, and editing. All AI-assisted content was critically reviewed, revised, and approved by the authors, who assume full responsibility for the accuracy, integrity, and originality of the work. No AI tool was used for data extraction, statistical analysis, or interpretation of results.

## RESULTS

### Study Selection

The PRISMA 2020 flow of records is summarised in Fig. 1. Database searches yielded 642 records; after removal of 200 duplicates and the inclusion of one additional record identified through hand-searching of reference lists, 443 unique records were screened at title and abstract level, of which 196 were excluded as not meeting the eligibility criteria. Following the priority-scoring step described in the Methods, 75 records were selected for full-text retrieval; nine of these could not be obtained despite the retrieval efforts detailed above, leaving 66 articles assessed in full-text eligibility.

**Fig. 1.**
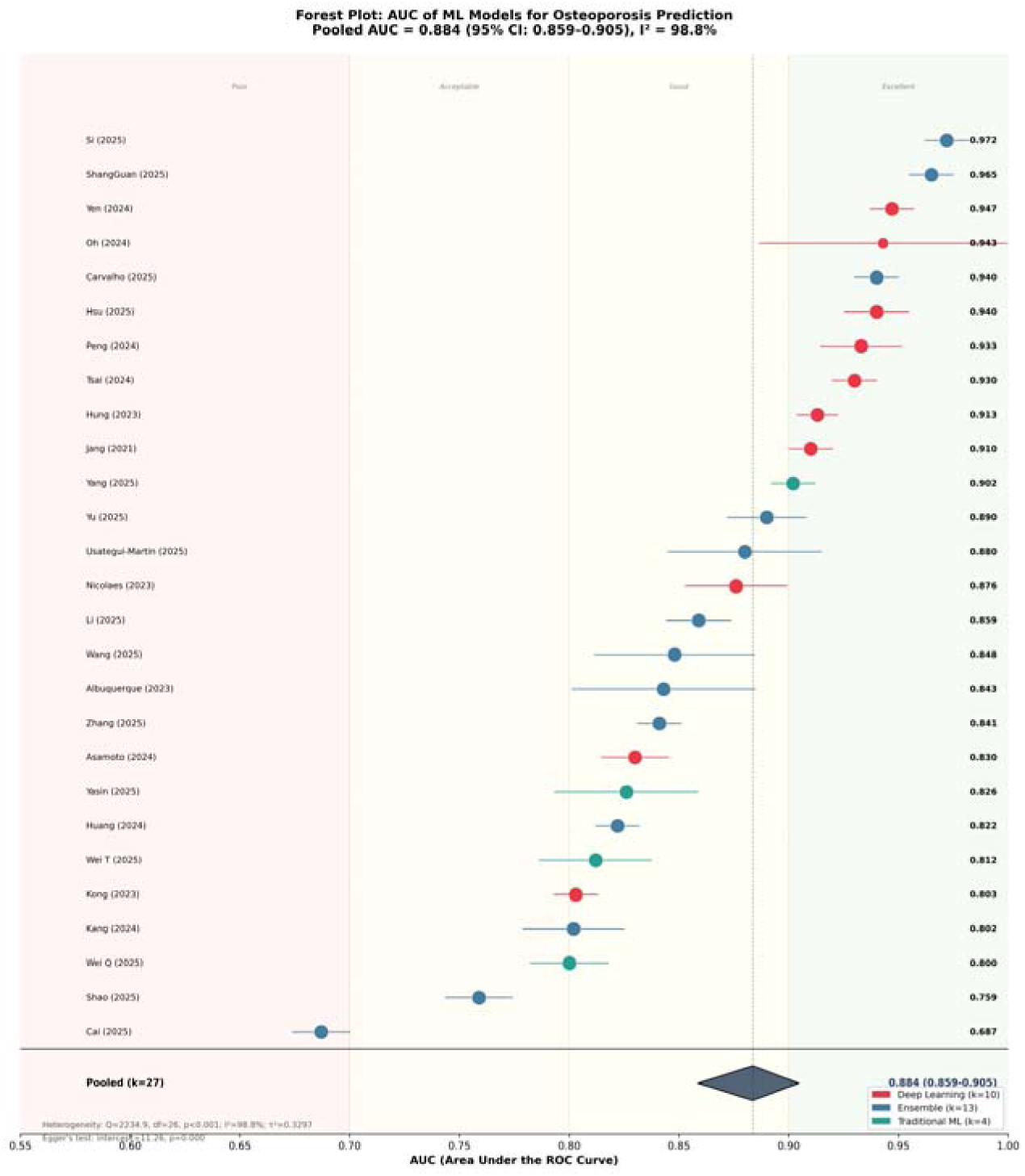
PRISMA 2020 flow diagram illustrating the identification, screening, eligibility assessment, and inclusion of studies. Records were identified from four electronic databases: PubMed (n = 298), Embase (n = 201), Web of Science (n = 87), and IEEE Xplore (n = 56), yielding 642 records in total. After removal of 200 duplicate records and addition of one manually identified record, 443 records were screened at the title and abstract level. Following full-text assessment of 66 records and PROBAST quality appraisal, 33 studies were included in the systematic review and 27 in the meta-analysis.

Full-text review led to the exclusion of 26 articles for the following reasons: not a prediction or classification model (n = 8), review article, editorial, or commentary (n = 5), outcome not related to osteoporosis or bone mineral density (n = 4), not using machine learning methodology (n = 3), insufficient methodological detail (n = 3), population outside scope (n = 2), and conference abstract only (n = 1). Four duplicate file versions were also identified and removed, yielding 36 unique studies for quality assessment.

PROBAST quality assessment was conducted on all 36 studies. During manual validation of studies with initial high or unclear risk ratings, three studies were excluded: one animal model study predicting bone mineral apposition rate in mice, one study predicting pharmacological treatment decisions rather than osteoporosis diagnosis, and one study classifying osteoarthritis from Raman spectroscopy data. The final systematic review included 33 studies (Fig. 1), of which 32 had retrievable full-text PDFs. Twenty-seven studies reporting area under the receiver operating characteristic curve (AUC) for classification tasks were included in the quantitative meta-analysis.

### Study Characteristics

The 33 included studies were published between 2021 and 2025, with the majority appearing in 2024 (n = 10, 30.3%) and 2025 (n = 15, 45.5%), reflecting the rapidly growing interest in this field. Studies originated from 14 countries, with China contributing the most publications (n = 9, 27.3%), followed by Taiwan (n = 6, 18.2%), South Korea (n = 4, 12.1%), and the United States (n = 2, 6.1%). Multinational collaborations accounted for five studies (15.2%), including contributions from Europe, East Asia, and the Americas. Study characteristics are summarized in Table 1.

Total sample sizes ranged from 112 to 50,114 participants (median = 5,328). The majority of studies employed retrospective cohort (n = 11, 33.3%) or cross-sectional (n = 10, 30.3%) designs, while three used prospective cohort data. Regarding population type, 15 studies (45.5%) enrolled general hospital or community populations, five (15.2%) focused on elderly individuals aged 50 years or older, three (9.1%) targeted postmenopausal women exclusively, and four (12.1%) studied disease-specific populations including patients with type 2 diabetes mellitus or chronic kidney disease.

Input data types varied considerably across studies. Twenty studies (60.6%) used clinical, demographic, or laboratory data, either alone or in combination. Nine studies (27.3%) used medical imaging as the primary input, including chest X-rays or radiographs (n = 4), computed tomography scans (n = 3), kidney-ureter-bladder radiographs (n = 1), and hand radiographs (n = 1). One study utilized a novel electromagnetic wave device, and another combined genomic with clinical data. Dual-energy X-ray absorptiometry served as the reference standard in 26 studies (81.8%), while three studies used quantitative computed tomography, two used ultrasound bone densitometry, and one relied on self-reported diagnosis.

### Machine Learning Methods and Validation Strategies

A wide range of machine learning algorithms were evaluated across the included studies (Table 2). Ensemble methods based on gradient boosting were the most frequently identified as best-performing models (n = 13, 39.4%), including XGBoost (n = 7), gradient boosting machines (n = 3), LightGBM (n = 1), CatBoost (n = 1), and a stacking ensemble (n = 1). Deep learning approaches were the second most common category (n = 12, 36.4%), comprising convolutional neural networks for imaging tasks (n = 8), artificial neural networks (n = 2), a U-Net architecture (n = 1), and a survival analysis model (DeepHit, n = 1). Random forest was the best model in five studies (15.2%). Logistic regression outperformed more complex algorithms in four studies (12.1%), notably in two studies employing interpretability-focused frameworks.

Most studies tested multiple algorithms for comparison. The median number of algorithms evaluated per study was four (range: 1–10). Feature selection methods were reported in 18 studies (54.5%), with LASSO regularization (n = 6), recursive feature elimination (n = 2), and Boruta (n = 2) being the most common approaches. The number of final input features ranged from 3 to 1,103, with a median of 11 features. Three studies used only three to five easily obtainable clinical variables, suggesting potential for simple screening tools.

Cross-validation was reported in 20 studies (60.6%), most commonly 10-fold (n = 9) or 5-fold (n = 4). Hyperparameter tuning was explicitly described in 22 studies (66.7%), primarily using grid search. Class imbalance handling was reported in only six studies (18.2%), with SMOTE and its variants being the most common approach (n = 4). External validation was performed in 14 studies (42.4%), using independent cohorts from different institutions, geographic regions, or national databases such as NHANES.

### Model Interpretability

Explainable artificial intelligence methods were employed in 20 studies (60.6%). SHAP (Shapley Additive Explanations) was the most common interpretability technique (n = 15), followed by LIME (Local Interpretable Model-agnostic Explanations, n = 4, often combined with SHAP), Grad-CAM for imaging models (n = 2), feature importance from tree-based models (n = 3), and attention-based visualization (n = 2). The most frequently identified top predictive features across clinical studies were age, body weight or body mass index, and alkaline phosphatase.

### Model Performance

Performance metrics of the best models from each study are presented in Table 3. Among the 27 classification studies reporting AUC, values ranged from 0.687 to 0.972 (median = 0.876, mean = 0.869). Twenty-two studies (81.5%) achieved an AUC above 0.80, and ten (37.0%) achieved an AUC above 0.90, indicating good to excellent discriminatory ability. Five studies were regression-based, reporting correlation coefficients (R = 0.68–0.961), R² values (0.218–0.841), or root mean square error as primary performance metrics.

Sensitivity was reported in 24 studies and ranged from 0.575 to 0.955 (median = 0.827), while specificity was reported in 24 studies and ranged from 0.511 to 0.973 (median = 0.799). Studies using imaging-based models tended to report higher sensitivity, consistent with the ability of deep learning to extract rich spatial features from radiographs and computed tomography scans. F1 scores, reported in 12 studies, ranged from 0.316 to 0.912 (median = 0.793). Calibration was assessed in 15 studies (45.5%), primarily through calibration curves, Brier scores, or decision curve analysis.

### Comparison with Existing Clinical Tools

Four studies directly compared their machine learning models with FRAX® or other established clinical tools. Zhang et al. (2025) [25] reported that gradient boosting (AUC = 0.841) significantly outperformed the Osteoporosis Self-assessment Tool (OST, AUC = 0.781, p < 0.0001) using only three features (weight, age, height). Kong et al. (2023) [26] demonstrated that DeepHit (AUC = 0.803) outperformed FRAX® (AUC = 0.691–0.716) for fracture prediction. In contrast, Usategui-Martín et al. (2025) [27] found that FRAX® (AUC = 0.92) slightly outperformed XGBoost (AUC = 0.88) for osteoporotic fracture prediction. Hung et al. (2023) [28] reported that an artificial neural network achieved 83.3% accuracy compared to 78.8% for FRAX®. Wang et al. (2025) [29] showed that XGBoost (AUC = 0.848) outperformed the OSTA tool (AUC = 0.739) in Tibetan women.

#### Meta-Analysis of Pooled Discriminative Performance

A random-effects meta-analysis using the DerSimonian–Laird method [30] was conducted on 27 studies reporting AUC for classification tasks. The pooled AUC was 0.879 (95% CI: 0.853–0.905), indicating good overall discriminative performance of machine learning models for osteoporosis prediction (Fig. 2). Substantial heterogeneity was observed (Q = 4577.2, I² = 99.5%, p < 0.001), which was expected given the diversity in study designs, populations, data types, and algorithms.

**Fig. 2.**
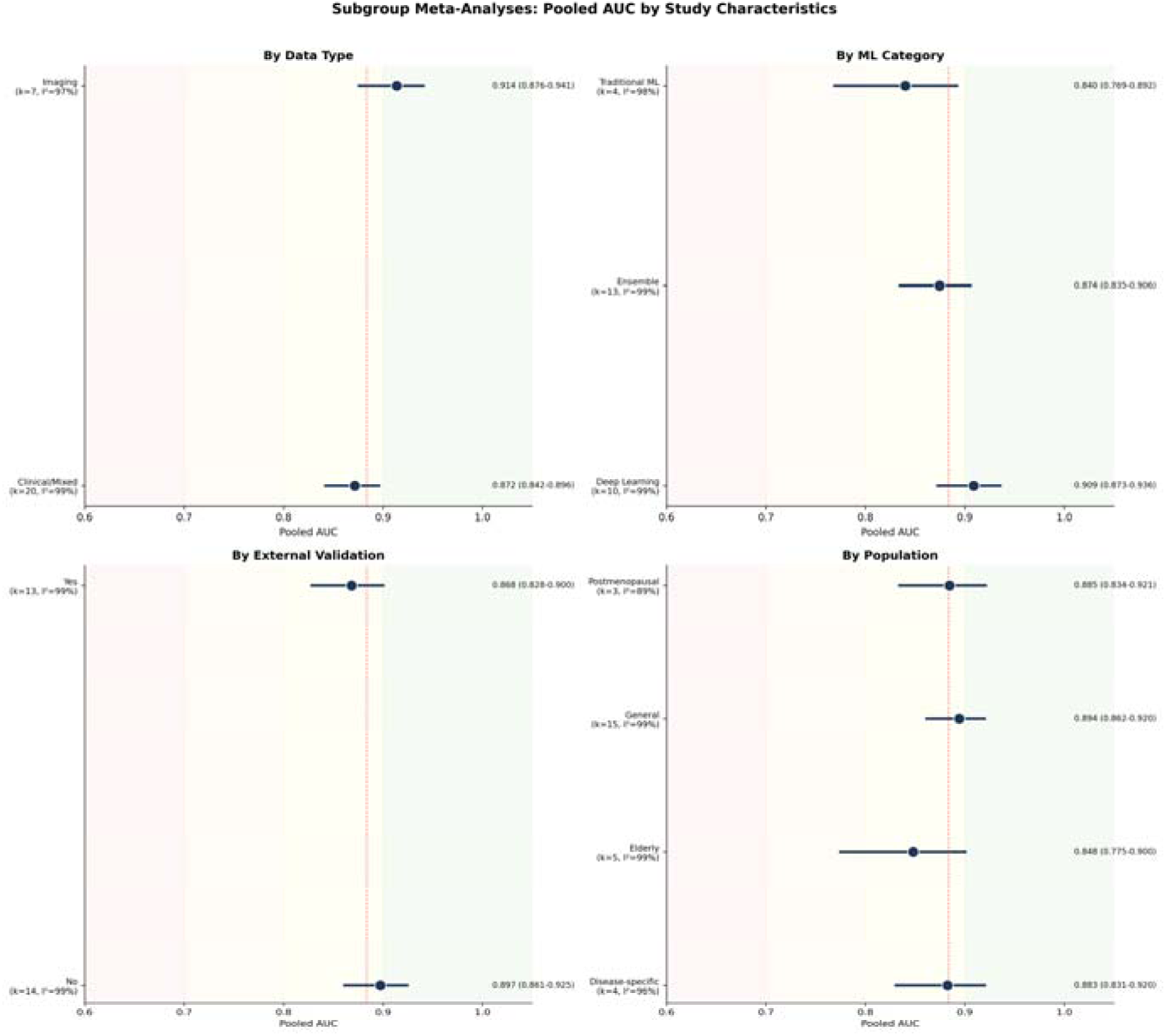
Forest plot of the area under the receiver operating characteristic curve (AUC) for machine learning models predicting osteoporosis (n = 27 studies). Individual study estimates are shown with 95% confidence intervals, color-coded by ML category: deep learning (red), ensemble methods (blue), and traditional ML (teal). The pooled AUC estimate from random-effects meta-analysis is shown as a diamond at the bottom (pooled AUC = 0.879; 95% CI: 0.853–0.905). Background shading indicates AUC quality zones: poor (<0.70), acceptable (0.70–0.80), good (0.80–0.90), and excellent (>0.90). Heterogeneity statistics (I² = 99.5%) and Egger’s test results are reported below the plot.

### Subgroup Analyses

Subgroup analyses were performed to explore sources of heterogeneity (Fig. 3). By data type, imaging-based models yielded a higher pooled AUC (0.905, 95% CI: 0.876–0.941, k = 7) compared to clinical or mixed-data models (0.872, 95% CI: 0.842–0.896, k = 20). By algorithm category, deep learning models achieved the highest pooled AUC (0.909, 95% CI: 0.873–0.936, k = 10), followed by ensemble methods (0.874, 95% CI: 0.835–0.906, k = 13) and traditional machine learning approaches (0.840, 95% CI: 0.769–0.892, k = 4).

**Fig. 3.**
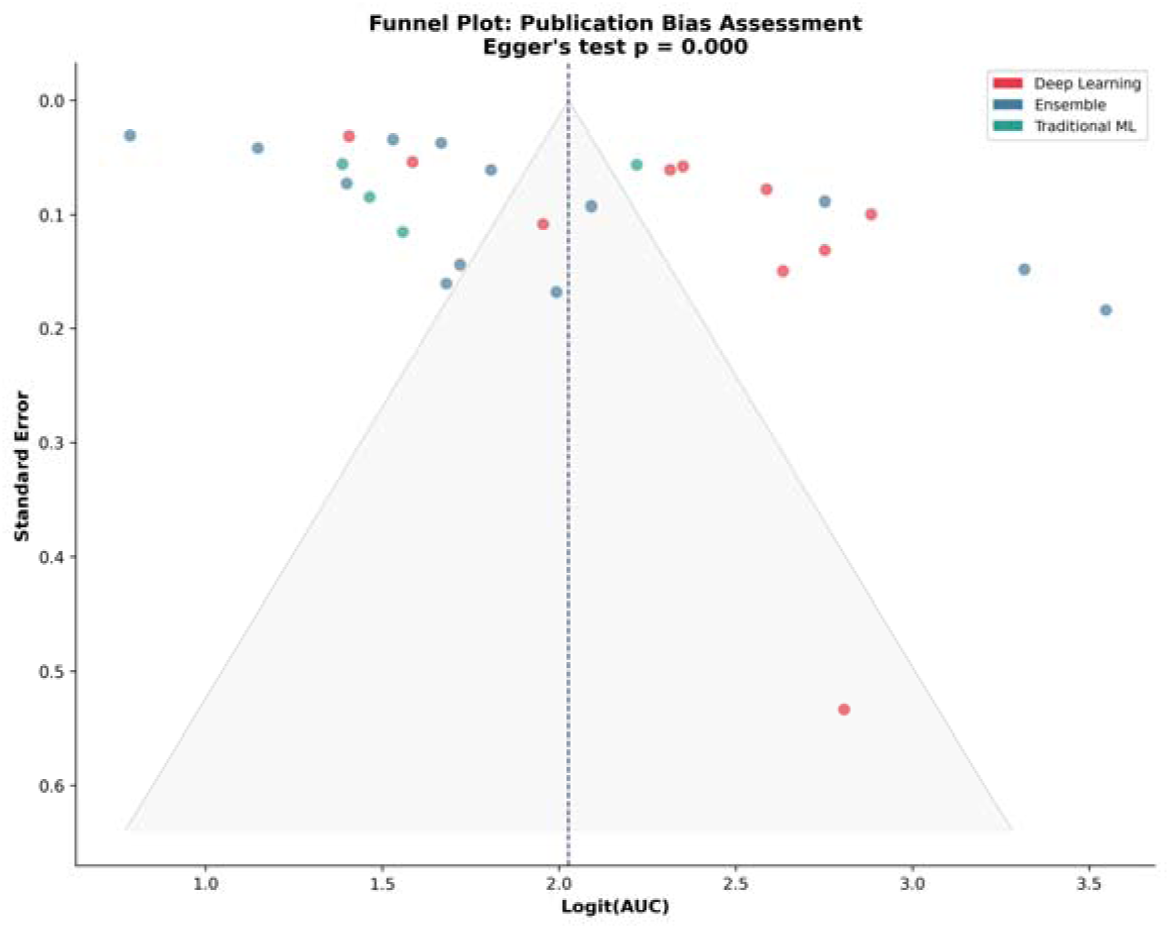
Subgroup analyses of pooled AUC by data type (clinical vs. imaging), machine learning category (deep learning, ensemble, traditional ML), external validation status, and population type. Boxes represent the pooled AUC estimate for each subgroup with 95% confidence intervals. Numbers in parentheses indicate the number of studies per subgroup.

Studies with external validation reported a pooled AUC of 0.868 (95% CI: 0.828–0.900, k = 13), which was lower than studies without external validation (0.897, 95% CI: 0.861–0.925, k = 14), suggesting a modest performance drop when models are tested on independent populations. By population type, studies in general populations achieved a pooled AUC of 0.894 (k = 15), postmenopausal women 0.885 (k = 3), disease-specific populations 0.883 (k = 4), and elderly populations 0.848 (k = 5). However, heterogeneity remained high within all subgroups (I² > 89%), indicating that other unmeasured factors contributed to performance variability.

### Publication Bias

Egger’s regression test revealed significant funnel plot asymmetry (intercept = 11.26, p < 0.001), suggesting potential publication bias or systematic differences between larger and smaller studies (Figure 4). Visual inspection of the funnel plot confirmed this asymmetry, with smaller studies tending to report higher AUC values. This finding should be interpreted with caution, as funnel plot asymmetry in diagnostic accuracy meta-analyses may reflect genuine heterogeneity rather than selective reporting, particularly given the diversity in methods, populations, and outcome definitions across the included studies.

**Fig. 4.** Funnel plot for assessment of publication bias among 27 studies included in the meta-analysis. Each point represents a study, plotted by logit-transformed AUC (x-axis) against standard error (y-axis, inverted). Points are color-coded by ML category: deep learning (red), ensemble methods (blue), and traditional ML (teal). The dashed vertical line indicates the pooled logit(AUC), and the gray triangle represents the 95% confidence interval pseudo-funnel. Asymmetry is evident, with smaller studies (higher SE) tending to report higher AUC values. Egger’s regression test confirmed significant asymmetry (intercept = 11.257, p < 0.001).

### Risk of Bias Assessment

Risk of bias was assessed using PROBAST for all 33 included studies. Overall, 32 studies (97.0%) were rated as low risk of bias, and one study (3.0%) was rated as unclear. No studies were classified as high risk. Three studies initially flagged as high or unclear risk were excluded during manual validation for fundamental methodological or scope issues (animal model, treatment prediction, and osteoarthritis classification). Among the four PROBAST domains, Domain 1 (Participants) and Domain 3 (Outcome) were generally well addressed, while Domain 4 (Analysis) was the most common source of concern, primarily due to incomplete reporting of calibration, class imbalance handling, or hyperparameter tuning details. This high proportion of low-risk studies should be interpreted cautiously: PROBAST criteria were developed primarily for traditional prediction models and may not fully capture sources of bias specific to machine learning, such as overfitting, data leakage, or inadequate handling of class imbalance.

Figure and Table References: Figure 1: PRISMA 2020 flow diagram; Table 1: Study characteristics; Table 2: ML methods and validation; Table 3: Performance metrics; Figure 2: Forest plot of pooled AUC; Figure 3: Subgroup analyses; Figure 4: Funnel plot.

## DISCUSSION

This systematic review and meta-analysis synthesises evidence from 33 machine learning studies published between 2021 and 2025, providing a structured appraisal of discriminative performance across diverse ML categories, data modalities, and validation strategies. The pooled AUC of 0.879 (95% CI: 0.853–0.905), derived from 27 classification studies, indicates that ML models can achieve clinically relevant discriminatory ability for osteoporosis prediction. This estimate must, however, be contextualised within extreme statistical heterogeneity (I² = 99.5%), evidence of publication bias favouring smaller studies, and a predominantly retrospective evidence base. These features collectively caution against interpreting the pooled AUC as a single generalizable benchmark; it is more appropriately understood as a descriptive summary of the performance range currently reported in a rapidly evolving and methodologically diverse field. Within these constraints, the findings support a potential complementary role for ML-based screening tools alongside established diagnostic pathways, particularly in settings where access to DXA is restricted.

The rapid growth in publications, with 45.5% of included studies published in 2025 alone, reflects the accelerating integration of artificial intelligence into musculoskeletal health research. This trend is likely driven by the convergence of increasingly available large-scale health databases (e.g., NHANES, UK Biobank), advances in deep learning architectures, and the growing recognition that osteoporosis remains substantially underdiagnosed worldwide. The World Health Organization has estimated that osteoporosis affects approximately 200 million individuals globally, yet screening rates remain low, particularly in low- and middle-income countries where DXA is not widely accessible [2].

### Principal Findings and Comparison with Previous Reviews

The pooled AUC of 0.879 observed in our meta-analysis exceeds estimates from two earlier systematic reviews: Smets et al. [15] reported a pooled AUC of 0.83 across 17 studies, while Amani et al. [16] reported 0.87 across 21 studies. This apparent improvement warrants cautious interpretation. In part, it may reflect genuine methodological advances, including the adoption of more complex model architectures, larger training datasets, and the growing uptake of external validation in recent years. However, it may equally reflect the well-documented tendency toward performance inflation in the machine learning literature, where hyperparameter optimisation and model selection routinely exploit test-set performance, and where publication bias selectively favours high-performing models [31]. The finding that externally validated studies yielded a lower pooled AUC (0.868; 95% CI: 0.828–0.900) than internally validated studies (0.897; 95% CI: 0.861–0.925) is consistent with this concern and underscores the importance of treating internally validated performance estimates with appropriate scepticism.

Our subgroup analyses revealed clinically meaningful differences across model categories. Deep learning models achieved the highest pooled AUC (0.909; 95% CI: 0.873–0.936), followed by ensemble methods (0.874; 95% CI: 0.835–0.906) and traditional ML algorithms (0.840; 95% CI: 0.769–0.892). The superior performance of deep learning models is particularly noteworthy, as these models were predominantly applied to imaging data, where convolutional neural networks can extract complex spatial features from radiographs and CT scans that are not readily captured by hand-crafted features. However, the clinical applicability of imaging-based deep learning models depends on the availability of the relevant imaging modality, which may limit their utility as first-line screening tools.

Imaging-based models showed higher pooled discriminative performance (AUC = 0.905; 95% CI: 0.876–0.941) compared to clinical/mixed data models (AUC = 0.872; 95% CI: 0.842–0.896). This pattern is consistent with the biological rationale that imaging modalities capture direct structural information about bone quality and density, whereas clinical models rely on indirect proxies such as age, body weight, and laboratory values. Nevertheless, the strong performance of clinical data–based models (AUC > 0.87) suggests that routinely available clinical variables can provide meaningful osteoporosis risk stratification without requiring additional imaging, which has important implications for population-level screening strategies.

### External Validation and Generalizability

An important finding of this review is the observed performance gap between internally and externally validated models. Studies without external validation reported a pooled AUC of 0.897 (95% CI: 0.861–0.925), compared to 0.868 (95% CI: 0.828–0.900) for externally validated models. Although this difference did not reach conventional statistical significance, the 2.9 percentage point reduction in AUC upon external validation suggests a degree of overfitting or dataset-specific optimization in models evaluated solely on internal test sets. This performance degradation is consistent with the broader machine learning literature on domain shift, where models trained on data from a specific population or institution may not fully generalize to new settings [32,33].

The proportion of studies performing external validation (42.4%) represents a notable improvement over earlier reviews but remains suboptimal. External validation is a critical step in the translational pathway from model development to clinical implementation, as it provides evidence that a model’s predictive performance is reproducible across different populations, time periods, and healthcare systems [20,31]. Future studies should prioritize multi-center external validation, ideally across diverse geographic and ethnic populations, to establish the robustness of ML-based osteoporosis prediction models. The use of standardized datasets and shared validation protocols, as advocated by the TRIPOD+AI [31] reporting guidelines, could facilitate more rigorous model assessment.

### Heterogeneity and Methodological Considerations

The substantial heterogeneity observed across studies (I² = 99.5%, Q = 4577.2, p < 0.001) warrants careful interpretation of the pooled estimates. High heterogeneity is common in meta-analyses of diagnostic and prognostic accuracy studies, where differences in study populations, outcome definitions, model architectures, and validation strategies contribute to between-study variability. In our review, studies varied substantially in sample size (112 to 50,114 participants), population characteristics (general vs. disease-specific), input data types (clinical vs. imaging), and ML algorithms employed. Although subgroup analyses partially explained this heterogeneity, particularly the data type and ML category subgroups, the residual I² within subgroups remained high (89.4–99.3%), suggesting that unmeasured sources of variation also contribute. While the pooled AUC of 0.879 indicates good overall discriminative performance, the extreme heterogeneity (I² = 99.5%) suggests that this estimate should not be interpreted as a single generalizable performance metric applicable across all settings, populations, or algorithmic approaches; rather, it reflects the broad range of achievable performance within a rapidly evolving and heterogeneous field.

The lack of standardization in study design and reporting represents a fundamental challenge in this field. Outcome definitions varied across studies, with some using the WHO T-score threshold of −2.5 for osteoporosis, while others used −1.0 for low BMD, and still others defined outcomes based on fracture occurrence. Similarly, reference standards differed: although 81.8% of studies used DXA as the gold standard, three used quantitative CT, two used ultrasound bone densitometry, and one relied on self-reported diagnosis. These inconsistencies in outcome definition and reference standard directly impact AUC estimates and contribute to the observed heterogeneity.

Several methodological limitations were identified across the included studies. Class imbalance was addressed in only 18.2% of studies, despite the typically unequal prevalence of osteoporosis and non-osteoporosis cases in study populations. Failure to account for class imbalance can lead to artificially inflated accuracy and AUC values, as models may achieve high performance simply by correctly classifying the majority class. Similarly, calibration, the agreement between predicted probabilities and observed outcomes, was assessed in only 45.5% of studies. Discrimination (AUC) alone is insufficient for clinical decision-making; a well-calibrated model is essential for informing individual patient management and establishing appropriate referral thresholds [34].

### Publication Bias

Egger’s regression test indicated significant funnel plot asymmetry (intercept = 11.257, p < 0.001), suggesting potential publication bias. Small-study effects were evident, with smaller studies tending to report higher AUC values. However, the interpretation of publication bias in diagnostic and prognostic accuracy meta-analyses requires caution. Funnel plot asymmetry can arise from genuine methodological differences between small and large studies rather than selective reporting alone. Smaller studies may use more homogeneous populations or highly curated datasets that favor higher performance, while larger studies, often using administrative or national health survey data, may include more heterogeneous populations with greater noise.

Additionally, the field of ML model development inherently involves a degree of selective reporting: researchers typically develop and compare multiple models, ultimately reporting the best-performing architecture. This practice, while standard in the field, can amplify apparent performance. The adoption of preregistration protocols and adherence to reporting guidelines such as TRIPOD (Transparent Reporting of a Multivariable Prediction Model for Individual Prognosis or Diagnosis) and its AI extension (TRIPOD+AI) [31] could mitigate these issues by promoting comprehensive reporting of all models tested, including negative results.

### Clinical Implications and Translational Potential

The findings of this review carry important clinical implications for osteoporosis screening and management. The strong discriminative performance of ML models based on routinely available clinical data (AUC = 0.872) suggests that these models could serve as effective pre-screening tools to identify individuals at high risk of osteoporosis who would benefit from confirmatory DXA scanning. Such a two-stage screening approach could improve the efficiency of osteoporosis detection programmes, particularly in primary-care settings where DXA access is limited. The clinical importance of this efficiency gain is amplified by the well-documented post-fracture care gap: international audits consistently estimate that approximately 80% of patients sustaining a fragility fracture are neither diagnosed with osteoporosis nor initiated on bone-protective therapy, despite their substantially increased risk of subsequent fracture [35]. Fracture Liaison Services (FLS), endorsed by the IOF Capture-the-Fracture initiative, have been shown to reduce this gap when systematically implemented, and ML-based pre-screening could plausibly extend their reach by enabling case-finding within unscreened primary-care populations [36]. This is especially relevant given that current clinical tools such as FRAX®, while validated and widely used, have recognized limitations, including moderate sensitivity and reliance on self-reported risk factors [12].

Four studies in our review directly compared ML models with FRAX® or other clinical tools, with mixed results. While some studies reported ML superiority, particularly when leveraging large feature sets or imaging data, others found that FRAX® performed comparably or even outperformed ML models for specific outcomes such as fracture prediction. These findings suggest that ML models and traditional clinical tools may serve complementary rather than competing roles. An integrated approach, in which ML models are used to enhance or supplement existing clinical workflows, may be more practical and effective than attempting to replace established tools entirely.

A particularly attractive translational paradigm is opportunistic osteoporosis screening, in which ML models are applied to existing computed-tomography (CT) examinations performed for unrelated clinical indications (e.g., abdominal CT for colorectal cancer screening or thoracic CT for lung-nodule surveillance) to derive vertebral attenuation values and infer bone status without exposing the patient to additional radiation or cost [37]. Several studies in the present review have applied deep-learning architectures to chest radiographs or routine CT slices in this paradigm, yielding pooled AUC estimates above 0.90 and offering a credible path to large-scale, low-marginal-cost case-finding in healthcare systems with high cross-sectional imaging volumes.

The adoption of explainable AI methods in 60.6% of studies is encouraging from a clinical implementation perspective. SHAP-based feature importance analyses consistently identified age, body weight or body mass index, and alkaline phosphatase as top predictive features, variables that are biologically plausible and clinically intuitive. This alignment between data-driven feature identification and established clinical knowledge enhances the face validity of ML models and may facilitate clinician acceptance. Three studies reported that models using as few as three to five easily obtainable variables achieved promising performance in development cohorts, suggesting potential for simple, deployable screening tools. These observations are consistent with broader work showing that SHAP-based feature attribution yields clinically coherent explanations across distinct musculoskeletal ML applications, including post-orthopaedic one-year mortality prediction from administrative and clinical variables [38] and osteoporosis risk prediction from anthropometric, demographic, and biochemical features in the NHANES cohort [38–40]. A complementary literature has further argued that model-dependent feature-importance rankings should be triangulated against threshold-free statistical measures of association (Spearman’s ρ, Kendall’s τ, mutual information, and total correlation) to mitigate the risk that apparent importance reflects algorithmic peculiarities rather than underlying biological structure [39].

Imaging-based deep learning models, particularly those utilizing chest radiographs or lumbar spine radiographs, offer an opportunistic screening paradigm. These models can retrospectively analyze radiographs obtained for other clinical indications, such as preoperative assessment or lung cancer screening, to identify patients with undiagnosed osteoporosis. Given that millions of radiographs are performed annually worldwide, opportunistic screening represents a scalable and cost-effective approach that does not require additional patient visits or imaging studies [41,42]. The pooled AUC of 0.905 for imaging-based models supports the feasibility of this approach.

### Risk of Bias Assessment

The PROBAST assessment yielded an unusually high proportion of low-risk ratings, with 32 of 33 included studies (97.0%) classified as low risk of bias. Several factors require consideration when interpreting this finding. First, the study selection process included a dedicated quality assessment stage during which three studies with fundamental methodological or scope issues were excluded, effectively raising the minimum quality threshold of the included evidence base. Second, and more importantly, PROBAST was designed primarily in the context of traditional regression-based prediction models [20] and may not adequately capture sources of bias that are particularly prevalent in machine learning research, including overfitting arising from insufficient sample sizes relative to model complexity, data leakage from shared preprocessing pipelines, and selective reporting of the best-performing algorithm from among many tested. The fact that class imbalance was addressed in only 18.2% of studies and that calibration was assessed in fewer than half of included studies are themselves indicators of methodological gaps that aggregate PROBAST ratings may have partially failed to detect, potentially leading to an overestimation of methodological quality in ML-based studies. Reviewers of this literature should therefore treat the overall quality profile with appropriate caution, and future systematic reviews in this domain may benefit from ML-specific extensions to existing risk of bias tools.

### Strengths and Limitations

This systematic review has several strengths. First, it provides the most comprehensive and up-to-date synthesis of ML models for osteoporosis prediction, including 33 studies published between 2021 and 2025, a period of rapid methodological advancement in the field. Second, the meta-analysis employed appropriate methods for pooling diagnostic accuracy measures, including logit transformation of AUC values and random-effects modeling to account for between-study heterogeneity. Third, extensive subgroup analyses by data type, ML category, external validation status, and population type provided nuanced insights into the sources of performance variation. Fourth, the PROBAST risk of bias assessment ensured that only methodologically sound studies contributed to the quantitative synthesis.

Several limitations should be acknowledged. First, the review protocol was not prospectively registered in a public registry such as PROSPERO, which limits the ability to assess potential deviations from the original analysis plan. Second, the high heterogeneity (I² = 99.5%) limits the interpretability of the pooled estimate, and the results should be viewed as a summary of the general range of performance rather than a precise point estimate. Third, the meta-analysis was restricted to studies reporting AUC, excluding five regression-based studies that used different performance metrics. Fourth, the SE of AUC was estimated using the Hanley–McNeil approximation for studies not reporting confidence intervals, which may introduce imprecision. Fifth, language restriction to English may have excluded relevant studies published in other languages, particularly Chinese, given the substantial contribution of Chinese research groups to this field.

### Future Directions

Several areas warrant attention in future research. First, prospective clinical validation studies are needed to assess the real-world impact of ML-based osteoporosis screening tools. While the retrospective evidence is promising, the translation from model development to clinical practice requires demonstration that ML predictions lead to improved patient outcomes, such as earlier diagnosis, timely treatment initiation, and fracture prevention. Randomized controlled trials or pragmatic clinical trials comparing ML-augmented screening pathways with standard care would provide the strongest evidence of clinical benefit.

Second, there is a need for greater standardization in study design and reporting. The adoption of the TRIPOD+AI guidelines [31], which provide specific recommendations for the reporting of prediction models involving artificial intelligence, would enhance the transparency, reproducibility, and comparability of future studies. Standardized outcome definitions (e.g., consistent use of WHO T-score thresholds), shared benchmark datasets, and common evaluation metrics would facilitate more meaningful cross-study comparisons.

Third, multimodal approaches that integrate clinical, imaging, genetic, and lifestyle data offer the potential for more accurate and personalized osteoporosis risk prediction. Only two studies in our review combined imaging with clinical data, and one study incorporated genomic information. Advances in data integration techniques and the increasing availability of electronic health records linking diverse data sources suggest that multimodal ML models may represent the next frontier in osteoporosis prediction.

Fourth, fairness and equity in ML-based screening deserve greater attention. Most studies in our review were conducted in East Asian or European populations, and the generalisability of these models to under-represented populations—including African, Latin American, South Asian, and Middle Eastern groups—remains uncertain. This gap is consequential because both BMD reference values and FRAX® calibration coefficients are population-specific, and indiscriminate application of a model trained in one population to another may introduce systematic miscalibration and inequitable case-finding. Given known differences in bone mineral density and fracture risk across ethnic groups [43], future studies should explicitly evaluate model performance across diverse populations and address potential algorithmic bias.

Finally, the integration of ML models into existing clinical workflows and electronic health record systems represents a critical implementation challenge. Technical barriers such as interoperability, model updating, and real-time inference must be addressed alongside clinical considerations such as appropriate referral thresholds, the role of ML predictions in shared decision-making, and medicolegal implications of algorithmic screening recommendations.

### Conclusions

In conclusion, this systematic review and meta-analysis suggests that machine learning models can achieve clinically relevant discriminative performance for osteoporosis prediction, with a pooled AUC of 0.879 across 27 classification studies. Deep learning models applied to imaging data and ensemble methods using clinical variables achieved the strongest subgroup estimates. These results are, however, substantially tempered by extreme heterogeneity (I² = 99.5%), evidence of publication bias, and a predominantly retrospective study design, which together preclude interpretation of the pooled estimate as a generalizable or clinically deployable performance benchmark. To realise the potential of ML-based osteoporosis screening, the field must advance from model development toward rigorous prospective validation, standardised outcome definitions, adherence to TRIPOD+AI reporting guidelines, and demonstrable evidence of real-world clinical impact.

## Data Availability

All data analysed in this systematic review and meta-analysis were extracted from previously published studies, which are cited in the manuscript. The complete data-extraction table, PROBAST risk-of-bias assessments, and summary tables are provided as supplementary materials.

## ACKNOWLEDGEMENTS

This study received Portuguese national funds from FCT - Foundation for Science and Technology through contracts UID/04326/2025, UID/PRR/04326/2025 and LA/P/0101/2020 (DOI:10.54499/LA/P/0101/2020).

## DECLARATIONS

### Conflict of interest

The authors declare that they have no conflict of interest.

### Ethics approval

This article does not contain any studies with human or animal participants performed by any of the authors. For this type of study formal consent is not required.

### Studies Included in Meta-Analysis

*The following citations correspond to the studies included in the systematic review and meta-analysis*.

| Citation | Title |
| --- | --- |
| [25] | A machine-learning-based osteoporosis screening tool integrating the Shapley Additive exPlanation (SHAP) method: model development and validation study |
| [26] | Development and validation of common data model-based fracture prediction model using machine learning algorithm |
| [27] | Assessment of the risk of osteoporotic bone fracture in postmenopausal women using machine learning methods |
| [28] | Establish and validate the reliability of predictive models in bone mineral density by deep learning as examination tool for women |
| [29] | Comparing machine learning models for osteoporosis prediction in Tibetan middle aged and elderly women |
| [40] | Enhancing osteoporosis risk prediction using machine learning: A holistic approach integrating biomarkers and clinical data |
| [44] | Osteoporosis screening using machine learning and electromagnetic waves. |
| [45] | External validation of a deep learning model for predicting bone mineral density on chest radiographs |
| [46] | Construction and validation of a multi-dimensional health indicator-driven osteoporosis risk prediction model: a large-sample cross-sectional study based on two centers |
| [47] | Machine learning models to predict osteoporosis in patients with chronic kidney disease stage 3-5 and end-stage kidney disease. |
| [48] | Development and validation of an explainable machine learning model for predicting osteoporosis in patients with type 2 diabetes mellitus |
| [41] | Opportunistic Osteoporosis Screening Using Chest Radiographs With Deep Learning: Development and External Validation With a Cohort Dataset |
| [49] | Construction of a novel online calculator for prediction of osteoporosis risk in Chinese type 2 diabetes patients |
| [50] | Towards Improved Identification of Vertebral Fractures in Routine Computed Tomography |
|  | (CT) Scans: Development and External Validation of a Machine Learning Algorithm |
| [42] | Evaluation of deep learning-based quantitative computed tomography for opportunistic osteoporosis screening. |
| [51] | A study on whether deep learning models based on CT images for bone density classification and prediction can be used for opportunistic osteoporosis screening. |
| [52] | Development of a machine learning-based predictive model for osteoporosis risk and its application in clinical decision support |
| [53] | Multidimensional dietary assessment and interpretable machine learning models predict the risk of prediabetes/diabetes and osteoporosis comorbidity in older adults |
| [54] | PrOsteoporosis: predicting osteoporosis risk using NHANES data and machine learning approach |
| [55] | Artificial Intelligence-enabled Chest X-ray Classifies Osteoporosis and Identifies Mortality Risk |
| [56] | Development and validation of an interpretable machine learning model for osteoporosis prediction using routine blood tests: a retrospective cohort study |
| [57] | Development and validation of machine learning models for osteoporosis prediction in chronic kidney disease patients: Data from National Health and Nutrition Examination survey |
| [58] | Machine learning-driven clinical decision support for low bone mineral density: A web-based prediction model with explainable AI integration |
| [59] | Predicting osteoporosis from kidney-ureter-bladder radiographs utilizing deep convolutional neural networks |
| [60] | HarDNet-based deep learning model for osteoporosis screening and bone mineral density inference from hand radiographs |
| [61] | Using Artificial Intelligence to Diagnose Osteoporotic Vertebral Fractures on Plain Radiographs |
| [62] | Interpretable machine learning model for low bone density screening in older adults using demographic and anthropometric data: findings from 2005 to 2020 NHANES |

## Notes

### Competing Interest Statement

The authors have declared no competing interest.

### Author Declarations

This is a systematic review and meta-analysis of previously published studies; no new human data were collected. All source data were extracted from peer-reviewed articles that were openly available before the initiation of this study, identified through PubMed, Embase, Web of Science, and IEEE Xplore. All included studies are cited in the manuscript, and the full data-extraction table is provided as supplementary material.

