## Supplemental Data 3 for "Machine Learning Models for Osteoporosis Prediction: A Systematic Review and Meta-Analysis"

### Databases and coverage

| Database | Interface / field tags | Records retrieved |
| --- | --- | --- |
| PubMed (MEDLINE) | Title/Abstract [tiab] | 298 |
| Embase | .ti,ab and Emtree terms | 201 |
| Web of Science Core Collection | Topic TS= | 87 |
| IEEE Xplore | “All Metadata” | 56 |
| **Total** |  | **642** |

Duplicates removed: 200 → **442 unique records**. One additional record identified by reference-list (citation) searching → **443 records screened**.

- **Publication date range:** 1 January 2020 – 28 February 2026.
- **Date last searched:** February 2026.
- **Language restriction:** English.
- **Filters applied:** publication date 2020–2026.

### Search queries

Twelve targeted queries, each combining a bone-health concept with a machine-learning / artificial-intelligence concept, were run in every database, adapting the field tags to each platform’s syntax. The queries are shown below in PubMed (Title/Abstract) syntax; the same concept terms were mapped to .ti,ab/Emtree in Embase, to TS= in Web of Science, and to “All Metadata” in IEEE Xplore. The publication-date filter 2020–2026 was applied to every query.

1. “machine learning”[tiab] AND “osteoporosis”[tiab] AND “prediction”[tiab]
2. “deep learning”[tiab] AND “osteoporosis”[tiab] AND (“screening”[tiab] OR “detection”[tiab])
3. “artificial intelligence”[tiab] AND “bone mineral density”[tiab]
4. “machine learning”[tiab] AND “fracture risk”[tiab] AND “prediction”[tiab]
5. “XGBoost”[tiab] AND “osteoporosis”[tiab]
6. “random forest”[tiab] AND “osteoporosis”[tiab] AND “prediction”[tiab]
7. “neural network”[tiab] AND “bone density”[tiab]
8. “FRAX”[tiab] AND “machine learning”[tiab]
9. “osteoporosis”[tiab] AND “NHANES”[tiab] AND “machine learning”[tiab]
10. “external validation”[tiab] AND “osteoporosis”[tiab] AND “prediction model”[tiab]
11. “ensemble learning”[tiab] AND “osteoporosis”[tiab]
12. (“explainable AI”[tiab] OR “explainable artificial intelligence”[tiab]) AND “osteoporosis”[tiab]

### Example — full PubMed strategy for query 1 (reproducible)

("machine learning"[Title/Abstract]) AND ("osteoporosis"[Title/Abstract]) AND
("prediction"[Title/Abstract]) AND ("2020/01/01"[Date - Publication] : "2026/02/28"[Date - Publication])

Eligibility screening, priority scoring, full-text assessment and reasons for exclusion are reported in the manuscript and summarised in the PRISMA 2020 flow diagram (Fig. 1).
